# Disease burden of liver cancer in China from 1990 to 2021 and its comparison with the global burden: insights from the Global Burden of Disease Study 2021

**DOI:** 10.1101/2025.03.30.25324914

**Authors:** Xinan Wan, Mingxing Fang, Haixia Diao, Le Yuan, Hang Zhang

**Affiliations:** Department of Oncology, The Second People’s Hospital of Wuhu, Wuhu, China

## Abstract

**Background:** China is a country with a very heavy burden of liver cancer disease, and understanding the epidemiological characteristics and trends of liver cancer in China can help develop targeted public health strategies.

**Methods:** The data were retrieved from the Global Burden of Disease (GBD) Study 2021. The age-standardized incidence rate (ASIR) and age-standardized death rate (ASDR) were used to estimate the trends in the incidence of and deaths from liver cancer by sex, region, country, and etiology between 1990 and 2021. Additionally, the attributable risk factors for deaths and disability adjusted life years (DALYs) were assessed. Finally, the Bayesian age-period-cohort model was used to predict the ASIR and ASDR of liver cancer from 2022 to 2035.

**Results:** In China, from 1990 to 2021, the incidence of liver cancer increased from 96.43 thousand (95% UI: 80.97, 113.76) to 196.63 thousand (95% UI: 158.27, 243.55), and the number of deaths increased from 94.93 thousand (95% UI: 79.88, 111.52) to 172.06 thousand (95% UI: 139.62, 212.49). Moreover, the ASIR was 10.58 (95% UI: 8.94, 12.43) per 100,000 population in 1990 and 9.52 (95% UI: 7.72, 11.78) per 100,000 population in 2021, and the ASDR was 10.75 (95% UI: 9.12, 12.61) per 100,000 population in 1990 and 8.35 (95% UI: 6.80, 10.29) per 100,000 population in 2021. The peak number of liver cancer cases was concentrated in the population aged 65-69 years, but the incidence rate showed an obvious trend of increasing with age. Liver cancer due to hepatitis B was the most common type of cancer. Tobacco ranked first among risk factors, with rapidly increasing risk factors being drug use and high body mass index. In the next 15 years, the ASIR and ASDR will not decrease among people aged 60 and over.

**Conclusion:** With the aging of the population in China, liver cancer remains an important disease burden. Therefore, measures should be taken to target risk factors and high-risk groups.

## Introduction

Liver cancer is the sixth most common malignant tumor and the third leading cause of cancer death after lung and colorectum cancer. Over three-quarters of the million liver cancer deaths worldwide occurred in 2022. This disease is the most common form of cancer death among males in some countries in Eastern Asia, Southeastern Asia, Northern and Western Africa, and Central America (1,2). In China, between 2005 and 2020, the mortality rate of liver cancer consistently ranked second among male cancer deaths, whereas among females, the mortality rate of liver cancer increased from third to second place (3). Liver cancer remains an essential worldwide public health problem. Therefore, it is necessary to analyze the burden of liver cancer in China and globally.

The Global Burden of Disease (GBD) Study 2021 included 371 diseases and injuries and 88 risk factors and combinations of risk factors for 204 countries and territories (4-6). This study aims to assess the global and Chinese burdens of liver cancer using data from the GBD Study 2021. We describe the global and Chinese epidemiological characteristics of liver cancer and identify variations in several factors (age, sex, and region). We also characterize the attributable risk factors for liver cancer. The incidence and mortality of liver cancer in China over the next 15 years are also predicted. By describing and predicting the burden of liver cancer in China and comparing it with the global situation, the development of a national-level strategy to reduce the disease burden caused by liver cancer in China can be promoted.

## Methods

The annual liver cancer incidence rates, deaths, and corresponding age-standardized rates by sex, region, country, and etiology between 1990 and 2021 were extracted from the GBD Study 2021 website (https://vizhub.healthdata.org/gbd-results/). Geographically, the world was divided into 21 GBD regions. Moreover, the 204 countries and territories were divided into five groups (low, low-medium, medium, high-medium, and high) according to the sociodemographic index (SDI).

The incidence rates of six types of liver cancer were evaluated. Additionally, the attributable risk factors for deaths and disability adjusted life years (DALYs) were assessed. Finally, we used the Bayesian age-period-cohort (BAPC) model (7,8) to predict the age-standardized incidence rate (ASIR) and age-standardized death rate (ASDR) of liver cancer from 2022 to 2035. All the statistical analyses and visualizations were conducted via the R statistical software program (version 4.1.2). A P value <0.05 was considered to indicate statistical significance.

Because the study was based on a publicly available dataset, this study was exempted by the ethics committee of the Second People’s Hospital of Wuhu city.

## Results

### Global burden and temporal trend in liver cancer

Globally, the number of incident cases of liver cancer increased from 244.68 thousand [95% uncertainty interval (UI): 224.79, 268.54] in 1990 to 529.20 thousand (95% UI: 480.33, 593.84) in 2021. In addition, the ASIR of liver cancer was 5.90 (95% UI: 5.43, 6.48) per 100,000 population in 1990 and 6.15 (95% UI: 5.58, 6.90) per 100,000 population in 2021. Moreover, the number of deaths caused by liver cancer worldwide was 483.87 thousand (95% UI: 440.40, 540.17) in 2021, increasing from 238.96 thousand (95% UI: 218.71, 263.03) in 1990. The ASDR of liver cancer was 5.86 (95% UI: 5.38, 6.46) per 100,000 population in 1990 and 5.65 (95% UI: 5.13, 6.30) per 100,000 population in 2021 (Table 1).

**Table 1:** Global incidence rates of and deaths from liver cancer in 1990 and 2021.

In China, the incidence of liver cancer increased from 96.43 thousand (95% UI: 80.97, 113.76) in 1990 to 196.63 thousand (95% UI: 158.27, 243.55) in 2021. Moreover, the number of liver cancer deaths increased from 94.93 thousand (95% UI: 79.88, 111.52) in 1990 to 172.06 thousand (95% UI: 139.62, 212.49) in 2021. The ASIR of liver cancer was 10.58 (95% UI: 8.94, 12.43) per 100,000 population in 1990 and 9.52 (95% UI: 7.72, 11.78) per 100,000 population in 2021, with an estimated annual percentage change (EAPC) of -0.27 [95% confidence interval (CI): -0.42, -0.13]. In addition, the ASDR of liver cancer was 10.75 (95% UI: 9.12, 12.61) per 100,000 population in 1990 and 8.35 (95% UI: 6.80, 10.29) per 100,000 population in 2021, with an EAPC of -0.76 (95% CI95% CI: -0.91, 0.61) (Table 1 and Figure 1).

**Figure 1.**
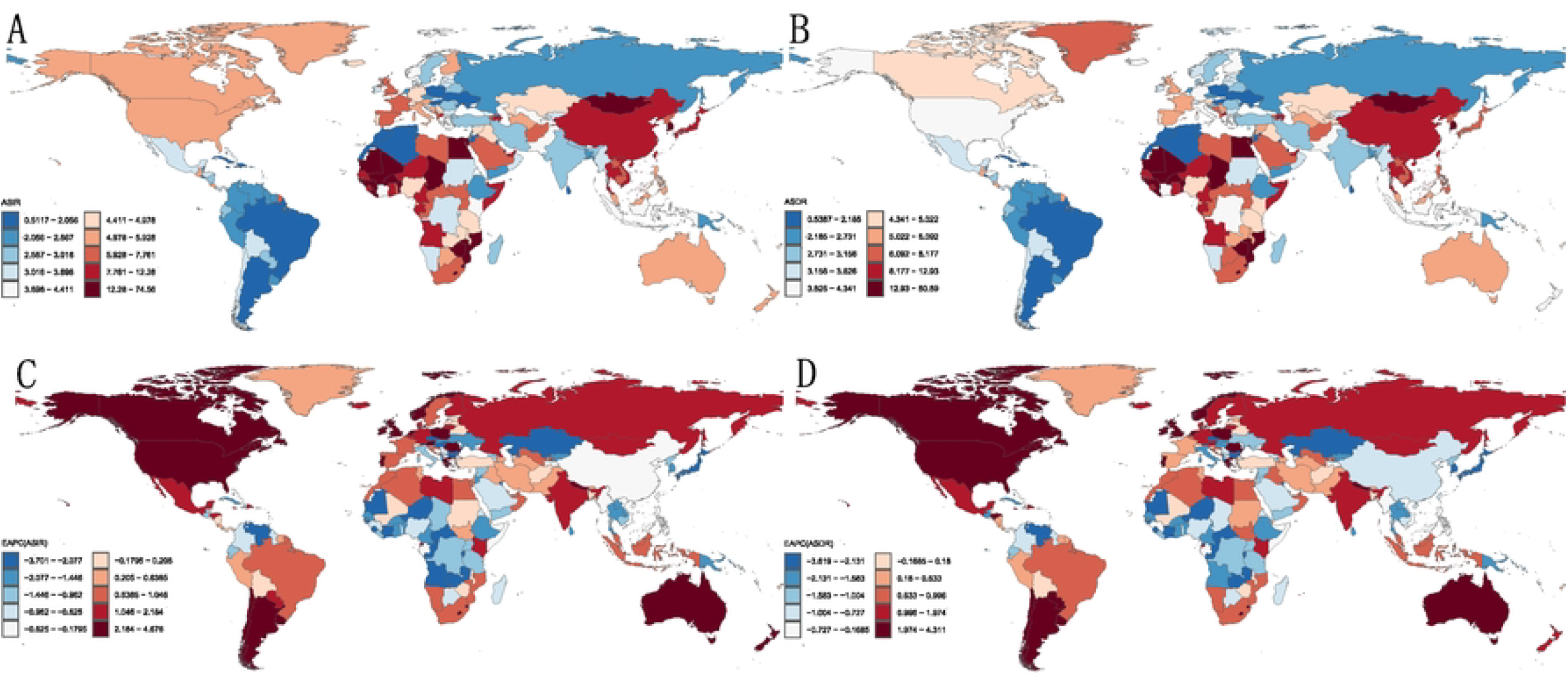
The ASIRs and ASDRs for liver cancer in 204 countries and territories. (A) The ASIR in 2021. (B) The ASDR in 2021. (C) The EAPC of the ASIR between 1990 and 2021. (D) The EAPC of the ASDR between 1990 and 2021.

Among the GBD regions, three high-income regions, namely, Asia Pacific, East Asia, and Western Sub-Saharan Africa, were among the regions with the highest ASIRs in 2021. In contrast, the lowest ASIRs in 2021 were observed in Southern Latin America, Tropical Latin America, and the Caribbean. Moreover, Western Sub-Saharan Africa, high-income Asia Pacific, and East Asia were the three regions with the highest ASDRs in 2021. Moreover, the lowest ASDRs in 2021 were observed in Southern Latin America, the Caribbean, and Tropical Latin America. Compared with those in 1990, the ASIRs in high SDI, high-middle SDI, and low-middle SDI regions increased. The ASIR in the other SDI regions showed a downward trend. The ASDRs in the high-middle SDI, middle SDI, and low SDI regions tended to decrease; in contrast, the ASDRs in the high SDI and low middle SDI regions increased (Table 1).

### Variation in liver cancer burden between males and females and among five-year age groups

In China, the ASIR was higher in males than in females in 2021. The number of incident cases in males was also greater than that in females in almost all age groups, except for the 90 years and above age group, which may be due to the larger population of females over 90 years old. The highest peak of liver cancer ASIRs occurred in the 90-94 years age group in males and the 85-89 years age group in females in 2021. The number of incident cases peaked in the 65-69 years age group for both sexes (Figure 2A). In GBD regions, a similar trend was observed, with males having higher ASIRs than females. The ASIRs of males and females were similar only in Andean Latin America (Figure 2B).

**Figure 2.**
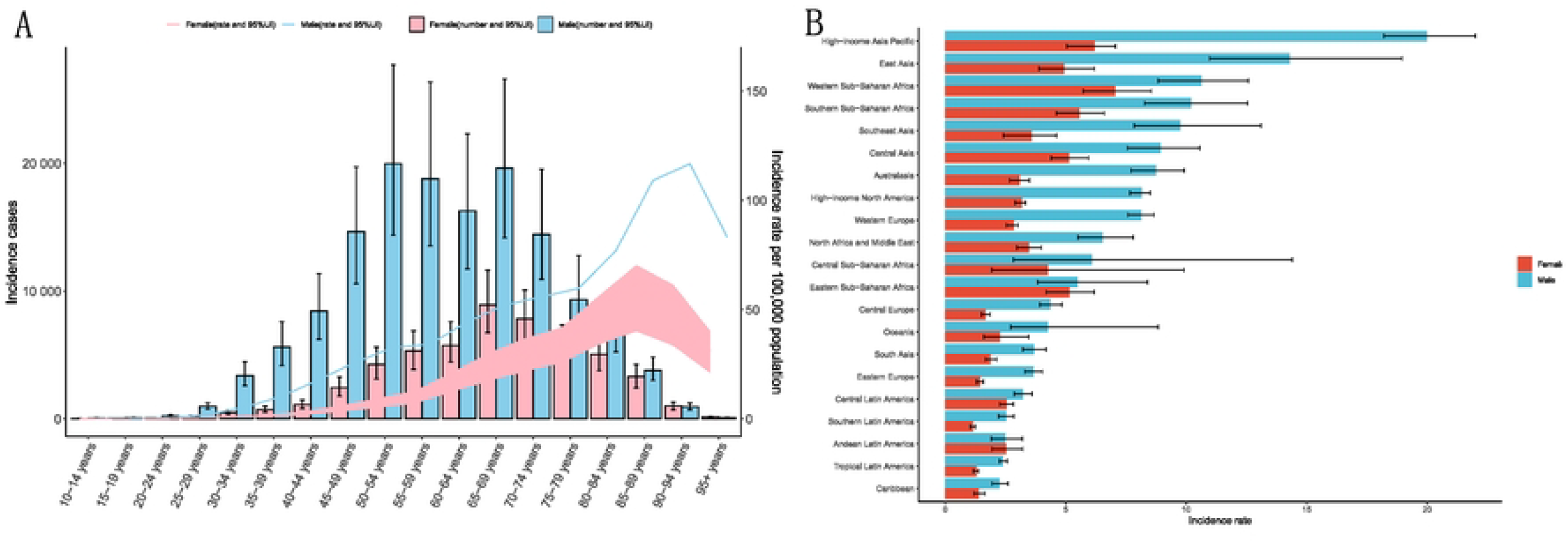
Total number and ASIRs by sex and age group in 2021. (A) Total number and ASIRs by sex and age group in China. (B) ASIRs by sex in GBD regions.

### Incidence rates of different types of liver cancer in China and at the regional level

In terms of etiology, there are six different types of liver cancer: liver cancer due to hepatitis B, liver cancer due to hepatitis C, liver cancer due to alcohol use, liver cancer due to NASH, hepatoblastoma, and liver cancer due to other causes. In China, liver cancer caused by hepatitis B had the highest ASIR in 2021 (5.73 per 100,000 population). The ASIRs of the other types were 1.77 per 100,000 population (liver cancer due to hepatitis C), 0.94 per 100,000 population (liver cancer due to alcohol use), 0.54 per 100,000 people (liver cancer due to NASH), and 0.08 per 100,000 population (hepatoblastoma).

In Central Asia, high-income Asia Pacific, high-income North America, Western Europe, Southern Latin America, Tropical Latin America, Central Latin America, North Africa and the Middle East, Eastern Sub-Saharan Africa, and Central Sub-Saharan Africa, the ASIR of liver cancer due to hepatitis C was the highest. In Eastern Europe, Central Europe, Australasia, and the Caribbean, the ASIR of liver cancer due to alcohol use was the highest (Figure 3).

**Figure 3.**
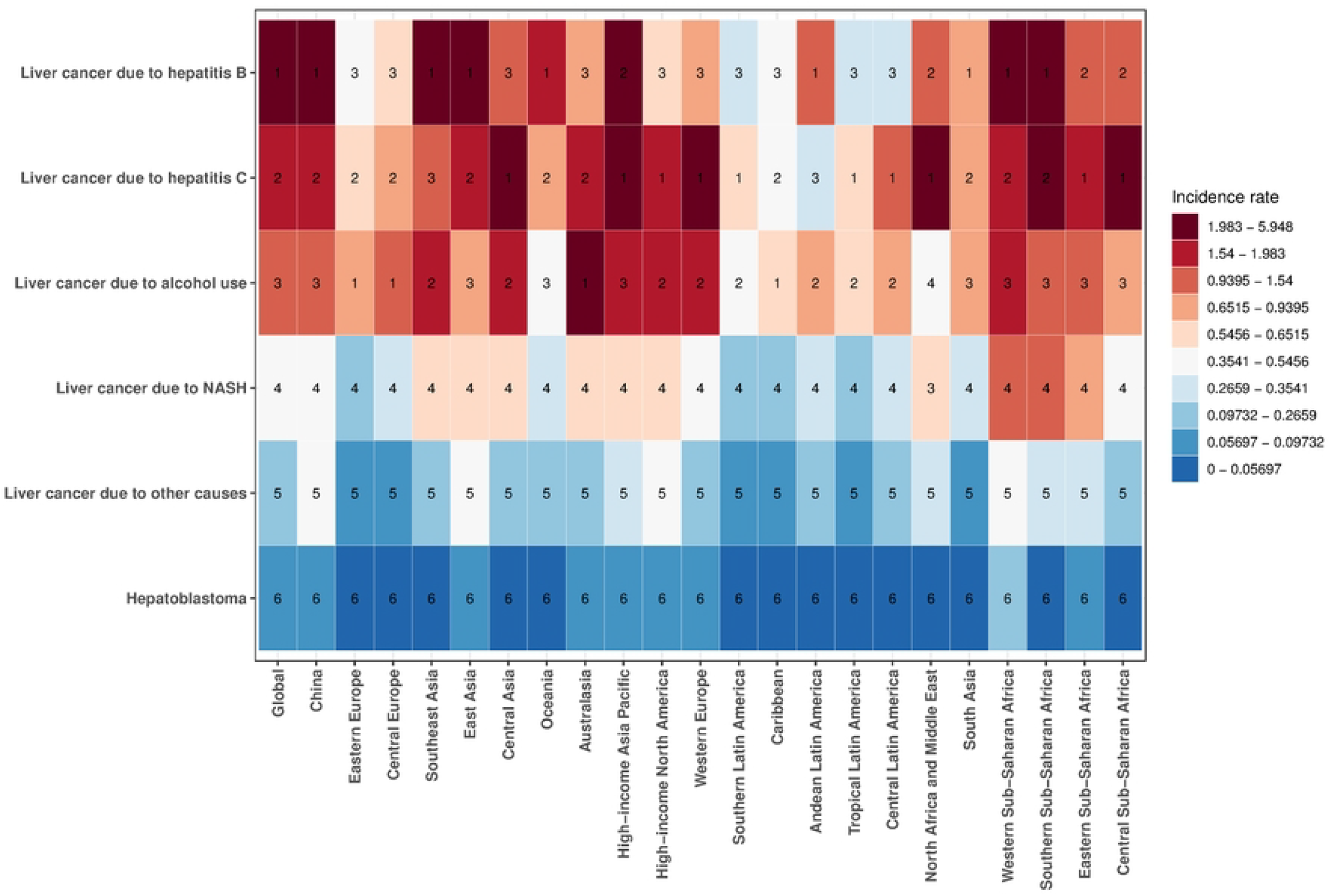
ASIRs of different types of liver cancer in 2021.

### Risk factors for liver cancer in China and at the regional level

We analyzed the attributable risk factors for deaths and DALYs in China from 1990 to 2021. According to the Global Burden of Disease Study 2021 framework for risk factor classification, risk factors associated with liver cancer were categorized into five levels. The level one risk factors include two types: behavioral risks and metabolic risks. The level two risk factors include five types: tobacco use, drug use, alcohol use, high body mass index, and high fasting plasma glucose. Among the risk factors associated with deaths, tobacco use ranked first. However, the fastest-growing risk factor was drug use, which rose from third place to second place, from 6.51% in 1990 to 11.55% in 2021. On the other hand, drug use and high body mass index increased rapidly among the risk factors attributable to DALYs, whereas the other three risk factors tended to be stable (Figure 4A, B).

**Figure 4.**
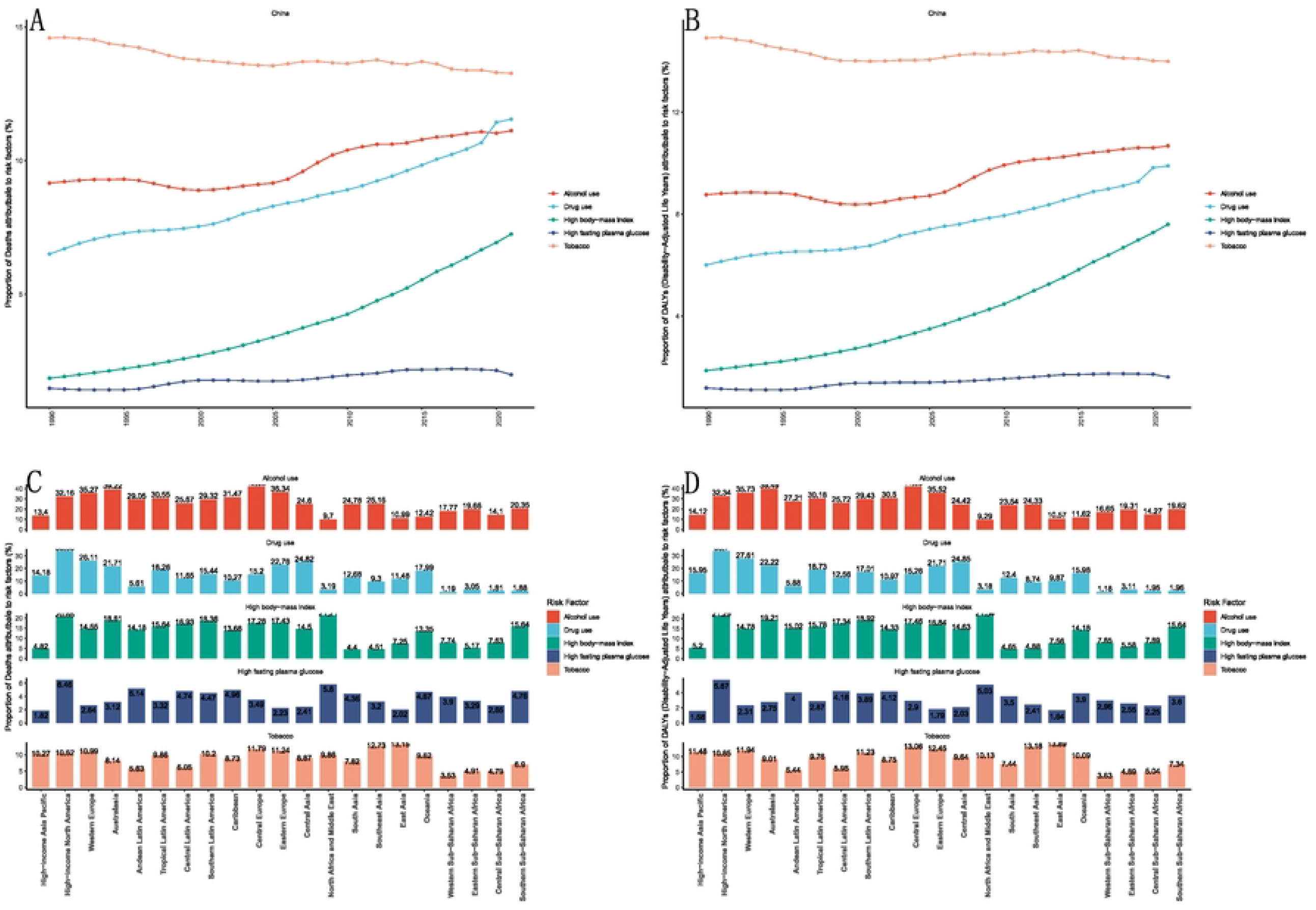
Deaths and DALYs attributable to risk factors (%). (A) Proportions of deaths attributable to risk factors in China from 1990 to 2021. (B) Proportions of DALYs attributable to risk factors in China from 1990 to 2021. (C) Proportions of deaths attributable to risk factors in GBD regions in 2021. (D) Proportions of DALYs attributable to risk factors in GBD regions in 2021.

Five secondary risk factors related to deaths were analyzed at the regional level in 2021. The three regions with the highest alcohol-attributable fractions were Central Europe (42.02%), Australasia (39.22%), and Eastern Europe (36.34%). For drug use, the highest fractions were in high-income North America (33.86%), Western Europe (26.11%), and Central Asia (24.82%). For high body-mass index, the highest fractions were in North Africa and the Middle East (21.21%), high-income North America (20.65%), and Australasia (18.61%). For high fasting plasma glucose, the highest fractions were in high-income North America (6.46%), North Africa and the Middle East (5.80%), and Andean Latin America (5.14%). For tobacco, the highest fractions were in East Asia (13.15%), Southeast Asia (12.73%), and Central Europe (11.79%). The five risk factors associated with DALYs were similarly distributed geographically (Figure 4C, D).

### Prediction of the liver cancer-related disease burden in China

According to research predictions, from 2022 to 2035, the ASIR and ASDR of liver cancer among the Chinese population will show a continuous downward trend (Figure 5A, B). The ASIR will decrease from 9.79 per 100,000 in 2022 to 8.73 per 100,000 in 2035. The ASDR will decrease from 8.66 per 100,000 in 2022 to 7.20 per 100,000 thousand in 2035. Further age group predictions revealed a significant downward trend in the ASIR and ASDR among the younger population, but stable trends in the population aged 60 and above (Figure 5C, D).

**Figure 5.**
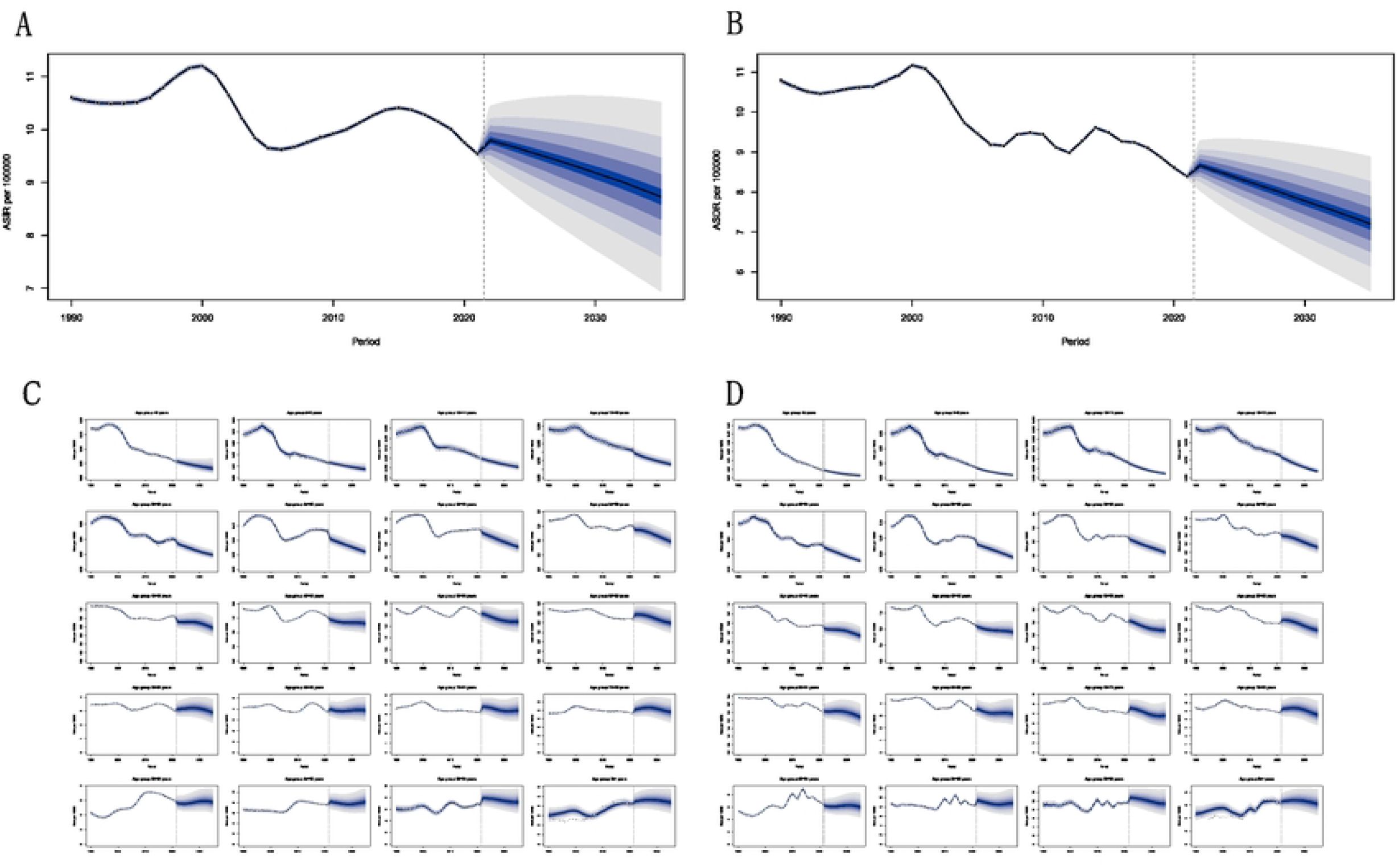
Predicted ASIR and ASDR of liver cancer in China in 2035. (A) The decreasing trend of the ASIR from 2022 to 2035. (B) The decreasing trend of the ASDR from 2022 to 2035. (C) The trend in the ASIR in different age groups. (D) The trend in the ASDR in different age groups.

## Discussion

China is a country with a very heavy burden of liver cancer, with the highest number of new cases and deaths from liver cancer in the world (9-11). From 1990 to 2021, although the number of new cases of liver cancer in China increased significantly, the ASIR decreased slightly from 10.58 per 100,000 population to 9.52 per 100,000 population. At the same time, the global situation is that the number of incident cases and the ASIR are increasing. In terms of the ASDR, both China and the world have experienced a decrease in the ASDR for liver cancer, but that in China is even greater, with EAPCs of -0.76 and -0.21, respectively. This reflects China’s initial achievements in the prevention and treatment of liver cancer. We found that the ASIRs of high-SDI, high-middle-SDI, and middle-SDI regions were greater than those of low-SDI regions, which may indicate that cancer diagnosis and reporting systems are more comprehensive in areas with higher social development indices.

Our findings reveal that the current peak in incident cases observed in China is focused on people aged 65-69 years. However, the peak liver cancer ASIR appears in males aged 90-94 years and females aged 85-89 years, and the incidence rate clearly tends to increase with age. With the aging of China’s population (12), the burden of liver cancer may gradually increase. In addition, the incidence rate of liver cancer in males is significantly higher than that in females, both in China and in most other regions of the world.

Liver cancer due to hepatitis B is the most common type of liver cancer in China. In 2021, its ASIR was 5.73 per 100,000 population, accounting for 60% of the total incidence rate of liver cancer (9.52 per 100,000 population). In addition, liver cancer due to hepatitis C was the most common type in Central Asia, high-income Asia Pacific, high-income North America, Western Europe, and other regions, indicating that viral hepatitis is the main cause of liver cancer (13-15).

The risk factors for liver cancer included five types: tobacco use, drug use, alcohol use, high body mass index, and high fasting plasma glucose (16-18). Tobacco use ranked first among risk factors, but its proportion tended to stabilize. The risk factors with rapid increases were drug use and high body mass index, which may be related to China’s rapid economic development in the past three decades and improvements in people’s material life. In other parts of the world, the risk factor alcohol use predominated, indicating that controlling tobacco and alcohol use is undoubtedly an effective measure to reduce the burden of liver cancer (19,20).

Furthermore, the projections indicate that the ASIR and ASDR of liver cancer in China will gradually decrease over the next 15 years. In terms of the predictions by age group, we found that the decrease in the ASIR and ASDR in the overall population was due to a decrease in the ASIR and ASDR among the younger population, whereas in the population aged 60 years and above, the ASIR and ASDR did not decrease. With the aging of China’s population, the high incidence and high mortality of liver cancer among the population over 60 is undoubtedly a serious disease burden.

## Limitations

However, our study had several limitations. First, the analysis heavily relied on the GBD database. Liver cancer diagnosis and reporting systems may not be sufficient in some underdeveloped countries and regions, leading to an underestimation of the true burden of liver cancer. Second, the GBD data does not include other risk factors for liver cancer, such as aflatoxin exposure (21). Third, we can only evaluate each etiology of liver cancer independently and cannot investigate the interactions between them.

## Conclusion

In China, the peak number of liver cancer cases is concentrated in the population aged 65-69 years, but the incidence rate clearly tends to increase with age. Liver cancer due to hepatitis B is the most common type of cancer in China, accounting for 60% of the total incidence rate of liver cancer. Tobacco use ranks first among the risk factors, with drug use and high body mass index being rapidly increasing risk factors. In the next 15 years, there will be no decrease in the ASIR and ASDR among people aged 60 and over. With the aging of the population in China, the high incidence rate and death rate of liver cancer among people aged 60 and over is undoubtedly a serious health problem. Therefore, liver cancer remains an important disease burden in China and the world, and it is necessary to take targeted measures on the basis of the epidemiological characteristics, common causes, and risk factors for liver cancer.

## Data Availability

The datasets generated and/or analyzed during the current study are available on the GBD Study 2021 website (https://vizhub.healthdata.org/gbd-results/). This public link to GBD database is open, and the use of data does not require additional consent from IHME.

## Acknowledgements

We thank all members of Global Burden of Disease Collaborative Network and Institute for Health Metrics and Evaluation (IHME).

## Author contributions

Conceptualization: Xinan Wan, Mingxing Fang

Data curation: Xinan Wan, Mingxing Fang, Haixia Diao

Formal analysis: Xinan Wan, Le Yuan, Hang Zhang

Methodology: Xinan Wan, Le Yuan, Hang Zhang

Software: Xinan Wan, Mingxing Fang

Supervision: Mingxing Fang

Visualization: Xinan Wan, Mingxing Fang

Writing - original draft: Xinan Wan, Mingxing Fang, Haixia Diao

Writing - review & editing: Xinan Wan, Mingxing Fang, Haixia Diao, Le Yuan, Hang Zhang

